# Increased Frequency of Acute Illness and Hospitalizations in Infants and Toddlers with Congenital Adrenal Hyperplasia

**DOI:** 10.1101/19005462

**Authors:** Teresa Tseng, Amy Seagroves, Christina M. Koppin, Madison Keenan, Elana Putterman, Eugene Nguyen, Sanjay Chand, Mitchell E. Geffner, Todd P. Chang, Mimi S. Kim

## Abstract

**Purpose:** Infants and toddlers with classical congenital adrenal hyperplasia (CAH) are at high risk for adrenal crisis and associated sequelae. To better understand acute illness at this early age, we determined the frequency and severity of acute illness and hospitalizations between 0-4 years of age, both within CAH and compared to controls. We also evaluated the impact of pre-hospital stress-dose hydrocortisone on Emergency Department (ED) visits and hospitalizations.

**Methods:** We performed a retrospective study of 40 CAH youth and 27 age-matched controls at a tertiary center. Characteristics of acute illnesses during the first 4 years of life were recorded, including fever, vomiting, diarrhea, ED visits, hospitalizations, abnormal electrolytes, and stress-dose hydrocortisone usage.

**Results:** CAH youth had more frequent illnesses requiring stress-dosing when they were younger than 2 years old [4.0 (1.0-6.0)] compared to when they were 2-4 years old [3.0 (1.0-4.0), P < 0.05], with the most illnesses during their first year of life. As well, CAH infants and toddlers had more hospitalizations younger than 2 years old compared to 2-4 years old (36 vs 2). 25% (3/12) of CAH youth with abnormal electrolytes in the ED did not receive any stress-dosing (oral/IM) prior to the ED, and only 25% (3/12) had received intramuscular hydrocortisone at home. CAH youth had more frequent ED visits (7.4 times as many) and hospitalizations (38 to 0) compared to controls.

**Conclusions:** Very young children with classical CAH are at high risk for acute illness and hospitalizations during their first 2 years of life, and do not receive adequate stress-dosing prior to the ED despite appropriate education. Our findings underscore the need for earlier recognition of acute illness in this vulnerable population and improved education regarding administration of stress-dose hydrocortisone to prevent morbidity.

## Introduction

Classical congenital adrenal hyperplasia (CAH) due to 21-hydroxylase deficiency is a potentially life-threatening form of primary adrenal insufficiency that affects ∼1:15,000 children, and is characterized by stress hormone (cortisol and epinephrine) and aldosterone deficiencies, along with androgen excess (1,2,3).

Infants and toddlers with CAH merit special attention, with increased all-cause mortality between ages 1 to 4 years, and a high risk for hypoglycemia, seizures, hospitalization, permanent cognitive impairment, and death caused by adrenal crisis during acute infectious illness (4,5,6,7). Newborns diagnosed with salt-wasting CAH require rapid replacement of cortisol, aldosterone, and salt to avoid catastrophic dehydration (6). In addition, newborns and infants with CAH exhibit epinephrine deficiency (2), adding risk for illness, adrenal crisis, and Emergency Department (ED) visits (8) at an age that inherently has an increased frequency of ED visits (9). Age has been noted to be an important factor in the longitudinal study of illness in CAH patients, with more illness events and stress-dosing noted to occur during childhood, and young age acting as a robust predictor of stress-dosing during this period of life (8). However, more needs to be understood about acute illness events and their pre-hospital treatment in infants and toddlers with classical CAH.

The main objective of this study was to characterize the frequency and severity of acute illnesses and hospitalizations in children with classical CAH during their infant and toddler years. We evaluated age, frequency of stress-dose illnesses and associated symptoms (*e*.*g*., fever ≥ 100.4°F, vomiting, and/or diarrhea), pre-hospital management of acute illness including stress-dose hydrocortisone, and severity of clinical presentation in the ED during the first 4 years of life. We hypothesized that the frequency and severity of illness would negatively correlate with age in CAH. We also compared CAH youth with age-matched controls, and hypothesized that there would be more frequent illness, ED visits, and hospital admissions in the CAH group compared to controls.

### Study Participants and Methods

This study was a retrospective chart review approved by the Children’s Hospital Los Angeles Institutional Review Board. Parents gave their written informed consent.

#### Study Participants

Forty children with classical CAH were studied from 1 month to 4 years of age (Table 1). CAH due to 21-hydroxylase deficiency was diagnosed biochemically based on a positive newborn screening test (California Department of Health Services, starting in 2005), and/or an elevated confirmatory serum 17-hydroxyprogesterone (17OHP) and androgen levels (testosterone and androstenedione) measured by liquid chromatography-tandem mass spectrometry LC-MS/MS (Esoterix, Calabasas Hills until 2009; Quest Nichols Diagnostics, San Juan Capistrano after 2009). Thirty-five percent of subjects underwent genetic *CYP21A2* analysis. There were 33 youth with the salt-wasting (SW) form and 7 with the simple-virilizing (SV) form; of the total, 65% (25/40) were female. Doses of glucocorticoid, mineralocorticoid, and sodium chloride were recorded for the children with CAH at both clinic and ED visits. CAH youth were on higher hydrocortisone (HC) and fludrocortisone doses during their first 3-6 months of life compared to when they were older, and so we excluded 0-6 month-old dose data from analyses of medication doses. Infants (with the exception of 3 SV) were treated with NaCl doses of 11.5 (9.7-15.8) mEq/day largely in the first year of life.

**Table 1.** Study Population.

| **Variable** | **N** | **Median (IQR)** |
| --- | --- | --- |
| **CAH** | **40** | **-** |
| **Sex** |  |  |
| Female | 26 (65.0%) | - |
| Male | 14 (35.0%) | - |
| **CAH Type** |  |  |
| Salt-wasting | 33 (82.5%) | - |
| Simple-virilizing | 7 (17.5%) | - |
| **Initial 17OHP (ng/dL)**<br>**[nmol/L]** |  |  |
| Newborn screen | 33 | 9260.0 (3736.9-17612.5)<br>[288.8 (135.1-537.6)] |
| Pre-treatment<br>confirmatory serum | 37 | 9875.0 (4931.5-25851.5)<br>[295.7 (147.6-790.1)] |
| Highest (newborn or<br>confirmatory) | 40 | 14423 (6465.0-26923.3)<br>[429.5 (189.2-822.0)] |
| **CONTROL** | **27** | **-** |
| **Sex** |  |  |
| Female | 20 (74.1%) | - |
| Male | 7 (25.9%) | - |

CAH youth were age-matched to 27 controls, with no significant difference in sex distribution between groups (Table 1). Controls had congenital hypothyroidism diagnosed by a positive newborn screen (elevated TSH, California Department of Health Services), but were otherwise healthy and euthyroid on replacement levothyroxine. The control group had a similar frequency of laboratory testing and follow-up visits as the CAH group.

Data from medical records were collected, spanning 20 years, including information that was recorded by a physician at clinic and ED visits at our institution. The number of acute illnesses, along with specific symptoms accompanying each illness (*i*.*e*., fever > 100.4° F, vomiting, and diarrhea), were assessed. Hospital treatment per illness-event was assessed from ED and inpatient admission records. In children with CAH, pre-hospital care was also assessed per illness-event, including HC stress doses administered during illness (stress-dose illness) and the route of administration [oral (PO) or intramuscular (IM) injection], as reported by caregivers. Pre-hospital transport was assessed, including administration of IM HC by Emergency Medical Services (EMS) personnel. ED serum electrolyte and glucose results were recorded, with electrolyte derangements deemed as a combination of one or more of the following: hyponatremia, hyperkalemia, and metabolic acidosis (low bicarbonate). Hypoglycemia was defined as glucose < 60 mg/dL (3.3 mmol/L). Interventions performed in the ED, such as administration of parenteral HC stress doses or intravenous fluids, were recorded from ED records.

#### Statistical Analyses

SPSS software version 24 (SPSS Inc., Chicago, IL) was used for all statistical analyses. Data were categorized into age groups of < 2 and ≥ 2 years old to evaluate modifying effects of age on illness frequency, and were normalized by person-year. Wilcoxon-Signed Rank tests were used for these comparisons. Chi-square tests and Spearman correlations were used for comparison of acute illness characteristics within the CAH group. Chi-square tests (for association) and Mann-Whitney U tests were used for comparisons between CAH and control youth. All statistical assumptions were investigated before assessment and addressed when necessary. The criterion for significance was α = 0.05 in all analyses, while the Benjamini-Hochberg procedure was used to control for false discovery rates. Results are shown as frequencies and percentages, or median and interquartile range (IQR).

## Results

### Illness Characteristics in CAH Youth

CAH youth had more frequent stress-dose illnesses when they were younger than 2 years old [4.0 (1.0-6.0)] compared to when they were 2-4 years old, per person-year [3.0 (1.0-4.0), Z = −2.37, P < 0.05; Figure 1]. In addition, CAH youth had more frequent stress-dose illnesses during their first year of life [3.0 (1.0-4.0)] compared to their second year of life [1.0 (0-2.0), Z = −3.03, P < 0.01].

**Figure 1.**
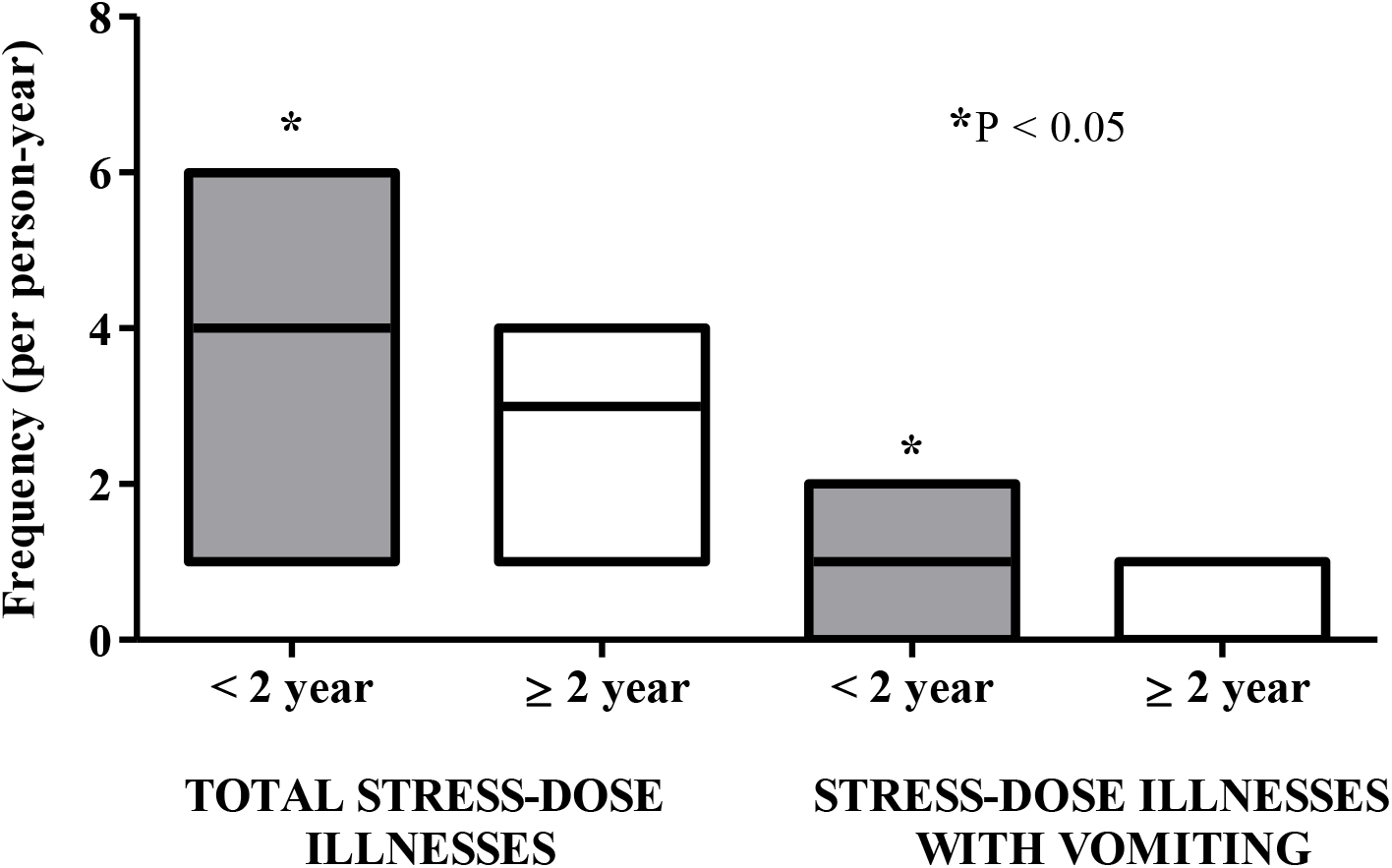
Illness characteristics in youth with classical CAH when they are younger than 2 years old and 2-4 years old. CAH youth had more frequent stress-dose illnesses when they were younger than 2 years old [4.0 (1.0-6.0)] compared to when they were ≥ 2 years old, per person-year [3.0 (1.0-4.0), P < 0.05]. CAH youth also had more frequent stress-dose illnesses involving vomiting between the ages of 0-2 years old [1.0 (0-2.0)] compared to ≥ 2 years old, per person-year [0 (0-1.0), P < 0.05].

CAH youth also had more frequent stress-dose illnesses involving vomiting when younger than 2 years old [1.0 (0-2.0)] compared to when they were 2-4 years old [0 (0-1.0), Z = - 2.20, P < 0.05; Figure 1]. However, there were no significant differences in frequency of stress-dose illnesses involving fever or diarrhea when the two age groups were compared.

### Pre-Hospital Maintenance Medications and Stress-Dosing

Maintenance medication doses were assessed over the first 4 years of life, excluding the first 6 months of life when doses could typically be higher. CAH youth exhibited a higher maintenance HC dose prior to 2 years old [15.1 (12.8-17.5) mg/m^2^/day] compared to 2-4 years old [13.5 (10.4-15.7) mg/m^2^/day; Z = −3.48, P < 0.001]. There was no difference in fludrocortisone dose prior to 2 years old and 2-4 years old.

Fludrocortisone dose correlated positively with frequency of overall stress-dose illness in CAH youth prior to 2 years old (R = 0.33, P < 0.05), and with illnesses involving vomiting (R = 0.38, P < 0.05) in particular. During this same age period, HC doses correlated with illnesses involving diarrhea (R = 0.38, P < 0.05). When older (2-4 years old), HC doses also correlated with total number of febrile illnesses in CAH youth (R = 0.36, P < 0.05), and trended towards significance with abnormal electrolytes (R = 0.29, P = 0.07). There were no significant correlations between HC or sodium chloride doses and frequency of stress-dose illness or with illnesses involving vomiting.

### Stress-Dosing and ED Visits

CAH youth had more frequent ED visits per person-year prior to 2 years old [1.5 (0.8-3.5)] than when they were 2-4 years old, per person-year [0 (0-1.0), Z = −4.17, P < 0.001; Figure 2]. Illnesses requiring a pre-hospital HC stress dose, either PO and/or IM, directly correlated with the number of ED visits (R = 0.75, P < 0.01). The highest newborn 17OHP level (*i*.*e*., newborn screen or initial pre-treatment confirmatory serum level; Table 1) was positively associated with frequency of ED visits in CAH youth between the ages of 2-4 years (R = 0.33, P < 0.05), but not when they were younger than 2 years old.

**Figure 2.**
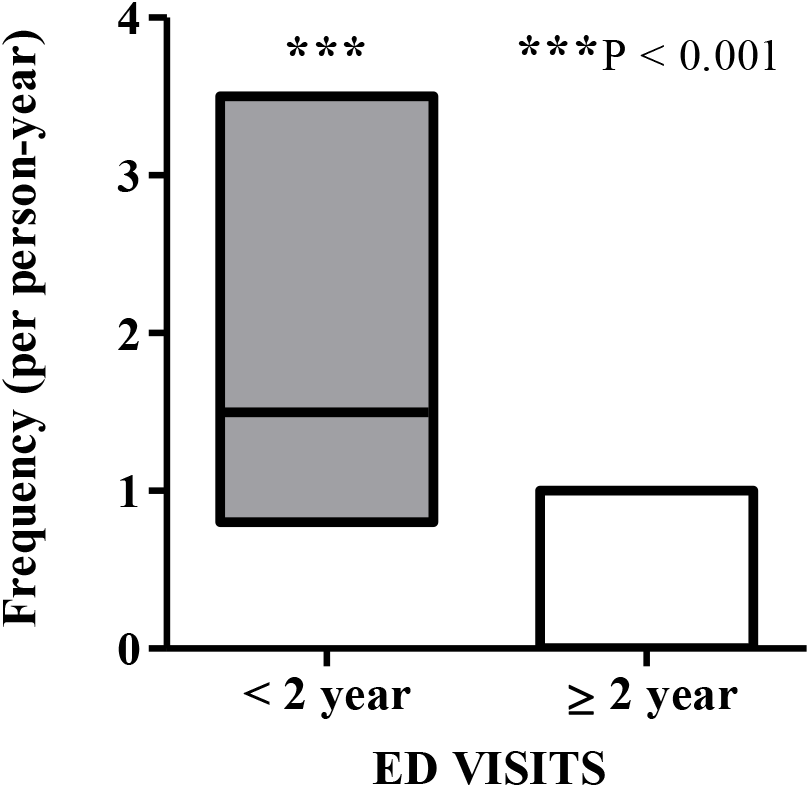
Emergency department visits in youth with classical CAH when they are younger than 2 years old and 2-4 years old. CAH youth had more frequent emergency department visits during acute illness when they were younger than 2 years old [1.5 (0.8-3.5)] compared to when they were 2-4 years old, per person-year [0 (0-1.0), P < 0.001].

Of 149 ED visits, there were 101 visits where CAH youth were given stress-dose HC (80 parenteral and 21 PO) in the ED. In regard to the former, 78.8% (63/80) of parenteral stress doses were administered in those who were younger than 2 years old and only 21.3% (17/80) were preceded by a PO stress dose given at home. Only 21.3% (17/80) of parenteral stress doses were administered when youth were between the ages of 2-4 years old.

Ten CAH youth received an IM HC injection at home and a subsequent intravenous (IV) HC stress dose in the ED. Two youth received an IM HC injection at home, but did not receive a parenteral stress dose when they were in the ED. Three youth received an IM HC injection at home, but were not brought to the ED afterwards.

### ED Visits, Abnormal Electrolytes and Hypoglycemia, and Hospitalizations

Electrolytes were checked in 74 of 149 ED encounters in CAH youth, with 29.7% (22/74) being abnormal. There were five times the abnormal electrolyte events in CAH infants and toddlers during their first 2 years of life (10 events) compared to when they were 2-4 years old (two events). Eight events occurred during infancy, and two events between 1-2 years of age. All 22 encounters of abnormal electrolytes in these age groups resulted in parenteral HC stress-dosing in the ED, of which 3 patients were subsequently admitted.

Only 25% (3/12) of infants and toddlers with abnormal electrolytes had received IM HC prior to the ED, with two of the three injections given in youth older than 2 years old. Half (6/12) of the cases had received a PO stress dose prior to the ED, but no IM HC, and 25% (3/12) had not received any stress-dosing (PO or IM) prior to the ED.

In addition, there were four events of hypoglycemia in infants and toddlers with CAH. Three of these events occurred during the first 2 years and one event occurred in a patient who was 2-4 years old. All four patients were PO stress-dosed prior to their hypoglycemic event and received parenteral HC at the ED. However, none of the patients had received IM HC prior to the ED.

Overall, when CAH youth were younger than 2 years old, they had 18 times as many hospitalizations compared to when they were 2-4 years old (36 vs 2). All 12 patients with abnormal electrolytes in the ED were hospitalized. Four ED encounters involving normal electrolytes also resulted in hospitalizations. One of these patients had received both PO and IM HC stress doses at home, followed by an additional IV HC stress dose in the ED. CAH youth did not have any hospitalizations requiring pediatric intensive care, nor were there any deaths.

### CAH vs Controls: Illness Characteristics

CAH youth between the ages of 0-4 years old had significantly more frequent total illnesses compared to controls [CAH 6.0 (3.2-9.0) vs controls 1.0 (0-4.0); Z = −5.19; P < 0.001], per person-year. As well, CAH youth had more frequent illnesses involving fever [CAH 2.6 (1.1-4.0) vs controls 0.5 (0-1.0); Z = −4.85; P < 0.001], vomiting [CAH 1.0 (0.8-2.8) vs controls 0 (0-0.5); Z = −4.70; P < 0.001], and diarrhea [CAH 1.0 (0-2.0) vs controls 0 (0-1.0); Z = −3.03; P < 0.01].

### CAH vs Controls: ED Visits and Hospitalizations

CAH youth between the ages of 0-4 years old had 7.4 times as many ED visits (149 visits in 36 cases) compared to controls (20 visits in 10 cases), with an overall increased median frequency of ED visits compared to controls [CAH 2.0 (1.0-4.8) vs controls 0 (0-1.0), Z = −4.30, P < 0.001]. CAH youth had a total of 38 hospitalizations, with 36 occurring during their first 2 years of life and only two between the ages of 2-4 years old. The control infants and toddlers did not have any hospitalizations.

## Discussion

This study demonstrates that infants and toddlers with classical CAH have a significant increase in frequency and severity of stress-dose illnesses during their first 2 years of life than when older. CAH youth have more illnesses associated with vomiting, ED visits, and hospitalizations when they are younger than 2 years old, compared to when they are 2-4 years old, carrying significance for an age group known to have a high risk for morbidity and mortality from adrenal crises (6,10). The adrenal stress-hormone deficiencies inherent to CAH (including epinephrine deficiency in infancy) likely add to this heightened risk of decompensation with acute illness generally seen in infants and toddlers (2,4,11). An even higher frequency of stress dose-requiring illness in the CAH youth that we studied occurred during their first year of life, further highlighting the risks at younger age in this condition.

We also found that infants and toddlers with CAH have more frequent ED visits and hospitalizations than age-matched controls, along with higher rates of illness and symptoms of fever, vomiting, and diarrhea. Morbidity associated with dehydration, hypoglycemia, and subsequent adrenal crisis is particularly concerning in very young children (12,13,14,15), leading us to consider the CAH infants and toddlers with abnormal electrolytes in the ED. The frequency of abnormal electrolytes and hypoglycemia was significantly higher when children were younger than 2 years old compared to between 2-4 years. This could also explain the correlation between fludrocortisone dose and stress-dose illness, with patients on higher daily fludrocortisone doses likely to have more severe salt-wasting during illness than patients on lower fludrocortisone doses. Our finding supports prior reports of fludrocortisone dose as a predictive marker for stress-dose requirement (8) and as a risk factor for hospitalizations in CAH (10).

The timely pre-hospital administration of stress-dose HC during acute illness seems critical to the prevention of an adrenal crisis and its associated morbidity and mortality (16). However, we found that only 25% (3/12) of CAH infants and toddlers with abnormal electrolytes in the ED had appropriately received a stress-dose injection of HC at home. In fact, 25% of the total cohort had not received any stress-dosing in the pre-hospital setting prior to the ED. It can be challenging for parents to administer IM HC despite having significant knowledge (17), perhaps due to technical difficulty with the kit or injection, lack of supplies, and/or psychological stress in a high-acuity situation involving a young child. Education in a multidisciplinary clinic could improve rates of IM HC administration during acute illness (18), although we still found reluctance to administer IM HC by parents at our center who were taught stress-dosing at diagnosis and had routine review at clinic visits by a clinician or nurse. We found that two emergently ill toddlers (extreme lethargy in one and a seizure in the other) also did not receive an injection from first responders, reflecting the lack of education regarding the emergency hydrocortisone injection in the state of California. This suggests that standardizing the EMS management of patients with adrenal insufficiency to include IM HC could minimize metabolic derangements and prevent major morbidity related to adrenal crises in very young children with CAH (19,20,21).

Despite most of our findings pointing towards an improvement in frequency and severity of illness in CAH youth with age, we did note a correlation between the frequency of ED visits and initial newborn 17OHP level when CAH youth were older toddlers (2-4 years old). Higher daily hydrocortisone dose also correlated with more febrile illnesses at this age. This suggests that CAH disease severity could continue to impact illness-related morbidity in older toddlers, despite the expectation that frequency and severity of illness would improve at this age. Only one-third of the sample had genetic testing, thus limiting analyses involving *CYP21A2* gene mutations, but additional markers of CAH severity would be useful to study further.

Our study was limited by the retrospective nature of data collection, in terms of available detail on illness-related measures such as the trajectory of symptoms, barriers to administration of critical pre-hospital stress dosing, and time to hospital intervention. The number of acute illness episodes may have been under-reported in controls, as illness history is not as critical a component of medical history for a child who does not have adrenal insufficiency. Thus, a prospective controlled study would help to better elucidate the connection between treatment during illness and clinical outcome, and a larger sample size would also allow for more in-depth analyses per year of life.

Our study highlights the heightened risk for acute illness, adrenal crisis, and related morbidity that exists in youth with classical CAH when they are younger than 4 years old, and even more so when they are under 2 years of age. Caregivers are rarely providing IM stress-dosing with HC at home in young children with CAH, especially in those cases found to have metabolic derangements in the ED, highlighting the need for better training of family caregivers and EMS personnel, to improve outcomes in children with classical CAH.

## Data Availability

The data availability via the primary investigator would require IRB approval for data-sharing.

## Acknowledgments

We are very grateful to the patients and families who participated in the Natural History Study of CAH from Infancy at our center. We thank CARES Foundation for support of the Children’s Hospital Los Angeles CAH Comprehensive Care Center, the Abell Foundation for ongoing support of CAH research at our center, and the Gustavus and Louise Pfeiffer Research Foundation Award for fellowship support. We also thank Heather Ross, Carol Winkelman, and Norma Martinez for administrative support of this project.

